# Acute and long-term impacts of COVID-19 on economic vulnerability: a population-based longitudinal study (COVIDENCE UK)

**DOI:** 10.1101/2022.03.03.22271835

**Authors:** Anne E Williamson, Florence Tydeman, Alec Miners, Kate Pyper, Adrian R Martineau

## Abstract

**Background:** Socio-economic deprivation is well recognised as a risk factor for developing COVID-19. However, the impact of COVID-19 on economic vulnerability has not previously been characterised.

**Objective:** To determine whether COVID-19 has a significant impact on adequacy of household income to meet basic needs (primary outcome) and work absence due to sickness (secondary outcome), both at the onset of illness (acutely) and subsequently (long-term).

**Design:** Multivariate mixed regression analysis of self-reported data from monthly on-line questionnaires, completed 1^st^ May 2020 to 28^th^ October 2021, adjusting for baseline characteristics including age, sex, socioeconomic status and self-rated health.

**Setting and Participants:** Participants (n=16,910) were UK residents aged 16 years or over participating in a national longitudinal study of COVID-19 (COVIDENCE UK).

**Results:** Incident COVID-19 was independently associated with increased odds of participants reporting household income as being inadequate to meet their basic needs, both acutely (adjusted odds ratio [aOR) 1.39, 95% confidence interval [CI] 1.12 to 1.73) and in the long-term (aOR 1.15, 95% CI 1.00 to 1.33). Exploratory analysis revealed the long-term association to be restricted to those who reported ‘long COVID’, defined as the presence of symptoms lasting more than 4 weeks after the acute episode (aOR 1.39, 95% CI 1.10 to 1.77). Incident COVID-19 associated with increased odds of reporting sickness absence from work in the long-term (aOR 5.29, 95% CI 2.76 to 10.10) but not acutely (aOR 1.34, 95% CI 0.52 to 3.49).

**Conclusions:** We demonstrate an independent association between COVID-19 and increased risk of economic vulnerability, both acutely and in the long-term. Taking these findings together with pre-existing research showing that socio-economic disadvantage increases the risk of developing COVID-19, this may generate a ‘vicious cycle’ of impaired health and poor economic outcomes.

**Trial registration:** NCT04330599

**Summary Box:** *What is already known on this topic:*

- Socioeconomic deprivation is recognised as a major risk factor for incidence and severity of COVID-19 disease, mediated via factors including increased occupational and household exposure to SARS-CoV-2 and greater physical vulnerability due to comorbidities
- The potential for COVID-19 to act as a cause, rather than a consequence, of economic vulnerability has not previously been characterised.

*What this study adds:*

- We demonstrate an independent association between incident COVID-19 and subsequent self-report of household income being inadequate to meet basic needs, both acutely and in the long term
- Incident COVID-19 was also associated with increased odds of subsequent self-report of sickness absence from work in the long-term.

## Introduction

The coronavirus disease 2019 (COVID-19) pandemic has threatened the health of the global population more than any crisis in living memory. Socioeconomic deprivation was recognised as a major risk factor for incidence and severity of disease prior to the development and roll-out of vaccination against severe acute respiratory syndrome coronavirus 2 (SARS-CoV-2), mediated via factors including increased occupational and household SARS-CoV-2 exposure and greater physical vulnerability due to comorbidities.^1,2,3^ This association persists in the vaccination era, with lower socioeconomic status associated with increased incidence and severity of breakthrough COVID-19.^4^ However, the potential for COVID-19 to act as a cause, rather than a consequence, of economic vulnerability has received less research attention, despite the fact that sustained symptoms following an acute episode (‘long COVID’) are common, with potential to impact negatively on people’s daily activities and capacity to work.^5^

One of the challenges in characterising effects of COVID-19 on economic well-being relates to the fact that that societal measures to control the spread of COVID-19 are detrimental to employment and economic participation, and may therefore have negative economic impacts even in those who do not experience disease themselves.^1,6^ Pre-pandemic analyses reveal a relationship between economic downturns and mortality, highlighting the risk of ‘deaths of despair’ arising from suicide, drug overdose, or alcoholism.^7^ The Brookings Institute draws a direct link between these vulnerable populations and the COVID-19 pandemic, with particular harms from COVID-19-related poverty observed among populations who are already vulnerable.^8^

In order to dissect out impacts of disease from the consequences of the societal response to the pandemic, we conducted a longitudinal population-based study that was initiated at the start of the pandemic, to determine whether incident COVID-19 was associated with subsequent markers of economic vulnerability. Our primary outcome was self-report of whether household income was sufficient to meet basic needs; this outcome captures individuals who consider themselves below the poverty line due to an adverse event.^9^ Our secondary outcome captured participants’ ability to participate in the workforce by asking whether individuals who developed COVID-19 were more likely to report absence from work due to sickness. Associations between incident COVID-19 and both outcomes were explored both acutely (i.e. at the time when a positive SARS-CoV-2 test result was reported) and subsequently (i.e. in the long term).

## Methods

### Study design, setting and participants

COVIDENCE UK is a prospective population-based cohort study of COVID-19 in the UK population.^10^ Its aims are to determine risk factors for incident COVID-19 in the UK population; to characterise the natural history of COVID-19 in the UK population; to evaluate the impact of COVID-19 on the physical, mental, and economic well-being of the UK population; and to provide a resource from which to identify potential participants for future clinical trials of interventions to prevent or treat acute respiratory infections. Inclusion criteria were age ≥16 years and UK residence at the point of enrolment. Participants were invited via a national media campaign to complete an online baseline questionnaire capturing COVID-19 status and a wide range of demographic, socioeconomic and clinical characteristics described below. Follow-up questionnaires at monthly intervals captured incidence of RT-PCR- or lateral flow test-confirmed SARS-CoV-2 infection, long-term symptoms of COVID-19 (‘long COVID’), and indicators of economic status. Specific questions are included in baseline and monthly questionnaires whose responses contributed data to the current analysis are displayed in Tables S1 and S2 of Supplementary Material. The study launched on 1^st^ May 2020, and this paper reports analyses of data collected up to 28^th^ October 2021. All participants who responded to the baseline questionnaire and provided data on SARS-CoV-2 test status and adequacy of household income to meet basic needs in at least one monthly follow-up questionnaire were eligible for inclusion in this analysis. Exclusion criteria for this analysis were self-report of a positive SARS-CoV-2 test, ‘long COVID’, or hospitalisation for COVID-19 prior to completion of the baseline questionnaire, and self-report of ‘long COVID’ in the absence of a positive RT-PCR or lateral flow test result for SARS-CoV-2.

### Definition of variables

Our primary outcome variable was self-report of a participant’s household income being insufficient to meet their basic needs. This was derived as a binary variable based on responses to the question: “Since you last checked in with us, has your household income been sufficient to cover the basic needs of your household, such as food and heating?”. Any answer other than ‘yes’ (namely: ‘no’, ‘sometimes’, or ‘mostly’) was coded as indicating insufficient income, whilst answering ‘yes’ was coded as indicating sufficient income. We also considered a secondary outcome associated with economic vulnerability, namely the ability to participate in the workforce. This was represented by a binary variable derived from responses to the question: “Which of the following best describes your current occupational status?”. Participants selecting ‘not working due to sickness, disability or illness’ from a drop-down menu were coded as being absent from work due to sickness.

The following covariates were selected prior to analysis based on their potential to act as confounders of the relationship between incident COVID-19 and study outcomes:^11^ age (classified as ‘working age’ [16-65 years] or ‘not working age’ [>65 years]), sex (male *vs* female, defined by sex assigned at birth), ethnicity (classified as white or minority ethnic origin), country of residence (England, Scotland, Wales, or Northern Ireland), Index of Multiple Deprivation (IMD) quartile of residential area,^12^ baseline occupational status (employed, self-employed, retired, furloughed, unemployed, student, never employed, not working due to sickness/disability/illness, or ‘other’), housing status (owns home outright, mortgage holder, private rental, renting from council, or other) and self-reported general health (poor, fair, good, very good, or excellent).

The principal independent variable of interest for our analysis main model was SARS-CoV-2 test positivity. This was defined by a binary indicator where ‘yes’ included any self-reported positive lateral flow or RT-PCR SARS-CoV-2 test result, and ‘no’ included either a self-reported negative lateral flow or RT-PCR SARS-CoV-2 test result or no report of any test taken. Associations between this variable and our two outcomes of interest were considered over two time periods. First, we built an ‘acute’ model to examine short-term effects of COVID-19 by asking whether SARS-CoV-2 test positivity was associated with increased risk of reporting insufficient income or sickness absence in the same month as the positive result was recorded. Second, we built a ‘long-term’ model to test whether the risk of reporting insufficient income or sickness absence was associated with a positive SARS-CoV-2 test result in that month or any month following.

We also conducted two exploratory analyses to determine whether there was a dose-response relationship for associations between COVID-19 severity and risk of reporting insufficient income. This was implemented by categorising participants reporting a positive SARS-CoV-2 test result according to their response to the question “Would YOU say that you currently have ‘long COVID’, i.e. ongoing symptoms more than four weeks after the onset of proven or suspected COVID-19?”. We compared those reporting ‘long COVID’ and those reporting a positive SARS-CoV-2 test result but no ‘long COVID’ to those without a positive SARS-CoV-2 test result (the referent category). Second, we categorised participants reporting a positive SARS-CoV-2 test result according to whether or not they were hospitalised, comparing those reporting hospitalisation for COVID-19 and those reporting COVID-19 not requiring hospitalisation to those without a positive SARS-CoV-2 test result (the referent category). Both of these exploratory analyses were conducted for the acute and long-term models as described above.

### Statistical analysis

Mixed effects logistic regression models were applied to assess the relationship between positive SARS-CoV-2 test results (RT-PCR or lateral flow) and reported insufficient income at any point prior to 28^th^ October 2021 in the main analysis. A random effect of unique participant identifier was included in all models to account for repeated measures, allowing assessment of within-participant variability. These analyses were adjusted for baseline socio-demographic characteristics as outlined above.

For analyses exploring potential impacts of ‘long COVID’ and disease precipitating hospitalisation, a random effect for a unique participant identifier was included to account for repeated measures, with adjustment for baseline characteristics as before, and substitution of the monthly-varying binary principal independent variable indicating SARS-CoV-2 test status with one of the other 3-level key independent variables as previously defined above (i.e. positive SARS-CoV-2 test result with subsequent ‘long COVID’, positive SARS-CoV-2 test result without ‘long COVID’ *vs* no positive SARS-CoV-2 test result, OR positive SARS-CoV-2 test result with hospitalisation, positive SARS-CoV-2 test result without hospitalisation *vs* no positive SARS-CoV-2 test result). The models including these 3-level variables were evaluated twice, firstly as standard categorical variables and then secondly exchanging categorical versions for numerical integers, which provided a p-value for trend for ‘long COVID’ and hospitalisation due to COVID-19 respectively, for both acute and long-term models. Models for each of these monthly-varying exploratory analyses were built separately from one another, and from the main model which categorised incident COVID-19 as a binary independent variable.

Mixed effects logistic regression models were also applied to assess the relationship between a positive SARS-CoV-2 test result and reported absence from work due to sickness at any point prior to 28^th^ October 2021. The insufficient income variable was not included in this secondary outcome model, and ‘long COVID’ and hospitalisation were also not considered. Missing data were assumed to be missing completely at random (MCAR) and were handled with listwise deletion in the generalised linear mixed models so that unbiased estimates were obtained. All statistical analyses were conducted using R version 4.1.1 with the mixed effects models conducted using R-package `lmer4.`

### Sub-group analyses

We tested for effect modification by including interaction terms for SARS-CoV-2 test positivity and age (categorised as ≤65 or >65 years) and sex (categorised as male or female at birth) in multivariable models investigating determinants of our primary outcome.

### Patient and public involvement

Three patient and public involvement representatives were involved in development of the research questions and the choice of outcome measures specified in the study protocol. One of them also led on development and implementation of strategies to maximise participant recruitment. Results of work will be disseminated to individual participants via a webinar.

## Results

19,980 participants completed the COVIDENCE UK baseline questionnaire between 1^st^ May 2020 and 29^th^ October 2021, of whom 1,412 did not complete any subsequent monthly questionnaire. Of the remaining 18,568 participants, 16,910 (91.2%) contributed data to the current analysis. Reasons for exclusion of the 1,658 participants who did not contribute data to this analysis are detailed in the participant flow diagram (Supplementary Appendix Figure S1). Table 1 presents baseline characteristics of participants contributing data to this analysis: their median age was 63 years, 69.8% were female, 94.7% were of White ethnic origin, 2.7% were receiving universal credit payments, 6.9% reported their household income as being ‘sometimes’, ‘mostly’ or ‘not’ sufficient to meet their basic needs in the month prior to enrolment, and 1.7% reported not working due to sickness. Figure 1 illustrates response flows in sufficiency of income to meet basic household needs over time.

**Table 1:** Participant characteristics at baseline.

|  |  | Number Participants (%) |
| --- | --- | --- |
| Sex | Male | 5106 (30.2%) |
|  | Female | 11,804 (69.8%) |
| Working age | Yes (16-65) | 10,338 (61.1%) |
|  | No | 6570 (38.9%) |
| Ethnicity | Minority ethnic | 894 (5.3%) |
|  | White | 16,015 (94.7%) |
| Country | Scotland | 1029 (6.1%) |
|  | Wales | 604 (3.6%) |
|  | Northern Ireland | 314 (1.9%) |
|  | England | 14,956 (88.4%) |
| IMD Quartile | 1 (most deprived) | 3990 (23.6%) |
|  | 2 | 4191 (24.8%) |
|  | 3 | 4299 (25.4%) |
|  | 4 (least deprived) | 4410 (26.1%) |
| Claiming universal credit | Yes | 464 (2.7%) |
|  | No | 16,390 (96.9%) |
| Occupation | Self-employed | 1554 (9.2%) |
|  | Retired | 7547 (44.6%) |
|  | Furloughed | 386 (2.3%) |
|  | Unemployed | 296 (1.8%) |
|  | Student | 345 (2.0%) |
|  | Other | 394 (2.3%) |
|  | Never Employed | 10 (0.01%) |
|  | Not working due to sickness | 281 (1.7%) |
|  | Employed | 6097 (36.1%) |
| Housing | Mortgage | 4250 (25.1%) |
|  | Private Renting | 1227 (7.3%) |
|  | Renting Council | 531 (3.1%) |
|  | Other | 724 (4.3%) |
|  | Owns home | 10,174 (60.2%) |
| Self-reported general health | Poor | 480 (2.8%) |
|  | Fair | 1808 (10.7%) |
|  | Good | 4537 (26.8%) |
|  | Very good | 6691 (39.6%) |
|  | Excellent | 3394 (20.1%) |
| Income sufficient to cover basic needs | Yes | 15,749 (93.1%) |
|  | Mostly | 617 (3.6%) |
|  | Sometimes | 147 (0.9%) |
|  | No | 396 (2.3%) |

1 Missing data: working age (N = 2, 0.01%), ethnicity (N = 1, <0.01%), country (N = 7, 0.04%), IMD quartile (N = 20, 0.12%), housing (N = 4, 0.02%), universal credit (N = 56, 0.3%), income sufficient (N = 1, <0.01%).

**Figure 1:**
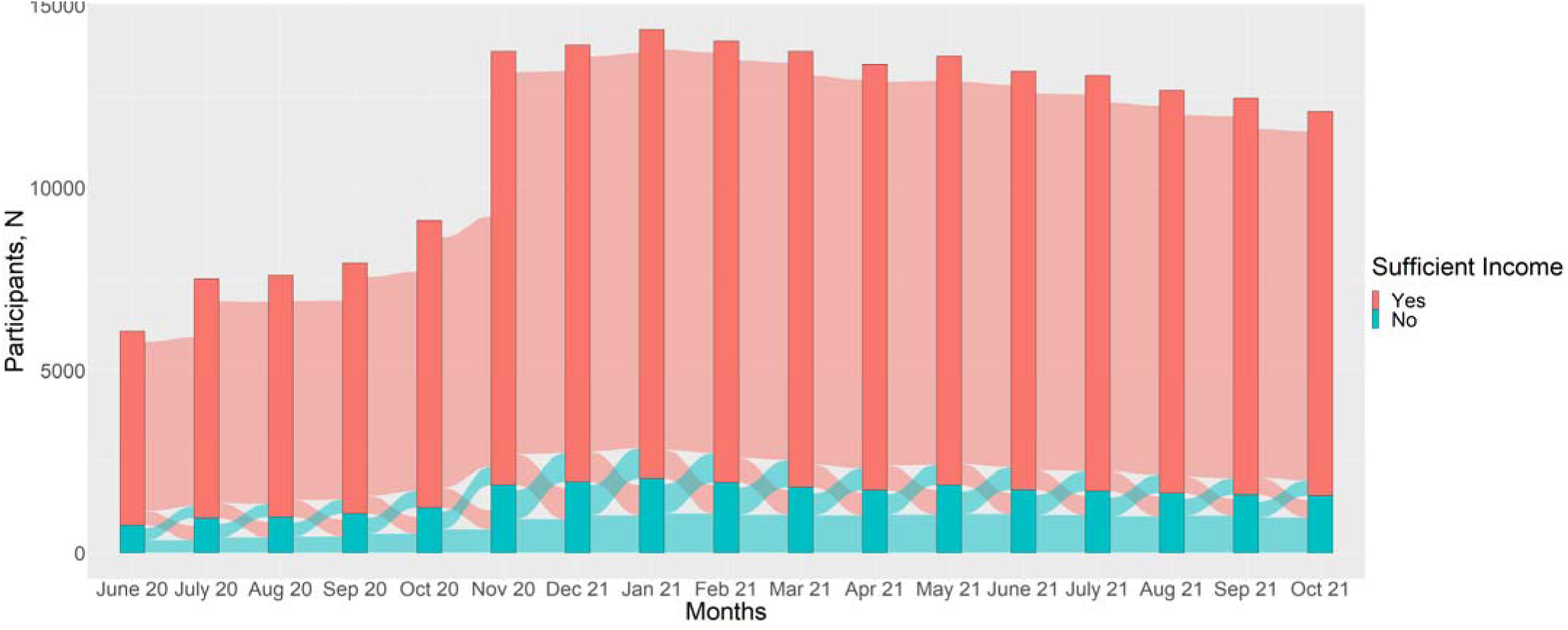
Sankey diagram illustrating response flows in sufficiency of income to meet basic household needs over time. Sufficient income was coded as ‘no’ when participants answered ‘no’, ‘sometimes’, or ‘mostly’ to the question, ‘Since you last checked in with us, has your household income been sufficient to cover the basic needs of your household, such as food and heating?’

A total of 1,120 participants reported a positive SARS-CoV-2 test result at least once between enrolment and the end of follow-up (28th October 2021). Of these, 39/1120 (3.5%) were hospitalised for COVID-19, and 308/1120 (27.5%) reported ‘long COVID’. A total of 7310/16,910 (43.3%) participants reported insufficient income on one or more occasions and 398/16,910 (2.4%) reported absence from work due to sickness on one or more occasions during follow-up (Supplementary Appendix Table S3).

Incident COVID-19 was independently associated with increased odds of participants reporting household income as being inadequate to meet their basic needs in multivariable analyses, both acutely (aOR 1.39, 95% CI 1.12 to 1.73) and in the long-term (aOR 1.15, 95% CI 1.00 to 1.33) (Table 2). Of the eight covariates included in each model, independent associations with increased risk of reporting insufficient income were also seen for non-White vs White ethnicity, younger vs older age (≤65 vs >65 years), higher vs lower deprivation quartile, poorer vs better health at baseline, being self-employed, furloughed, other (including sick) or unemployed vs being employed at baseline, and having a mortgage, privately renting, or renting from the council vs owning their home outright. Neither sex nor age modified the association between SARS-CoV-2 test-positivity and reporting insufficient income (for sex, P for interaction = 0.23 and 0.14 for acute and long-term models, respectively; for age, P for interaction =0.48 and 0.18 for acute and long-term models, respectively).

**Table 2:** Determinants of reporting insufficient income during follow-up.

| Variable | Response | Acute Responses |  | Long-term Responses |  |
| --- | --- | --- | --- | --- | --- |
|  |  | Adjusted OR (95% CI) | P | Adjusted OR (95% CI) | P |
| Incident COVID-19 | Yes | 1.39 (1.12 to 1.73) | 0.002 | 1.15 (1.00 to 1.33) | 0.049 |
|  | No | 1.00 | - | 1.00 | -- |
| Sex | Male | 1.03 (0.90 to 1.11) | 0.548 | 1.03 (0.93 to 1.15) | 0.549 |
|  | Female | 1.00 | - | 1.00 | - |
| Age, years | 16-65 | 1.63 (1.41 to 1.87) | <0.001 | 1.63 (1.41 to 1.87) | <0.001 |
|  | >65 | 1.00 | - | 1.00 | - |
| Ethnicity | Minority ethnic | 1.83 (1.49 to 2.27) | <0.001 | 1.83 (1.49 to 2.27) | <0.001 |
|  | White | 1.00 | - | 1.00 | - |
| Country | Scotland | 1.01 (0.80 to 1.28) | 0.916 | 1.02 (0.80 to 1.29) | 0.898 |
|  | Wales | 0.82 (0.61 to 1.09) | 0.164 | 0.82 (0.61 to 1.10) | 0.187 |
|  | Northern Ireland | 0.91 (0.62 to 1.31) | 0.588 | 0.91 (0.63 to 1.32) | 0.608 |
|  | England (ref) | 1.00 | - | 1.00 | - |
| IMD Quartile | 1 (most deprived) | 1.51 (1.27 to 1.79) | <0.001 | 1.50 (1.27 to 1.79) | <0.001 |
|  | 2 | 1.26 (1.10 to 1.44) | <0.001 | 1.26 (1.10 to 1.45) | <0.001 |
|  | 3 | 1.04 (0.91 to 1.20) | 0.535 | 1.05 (0.92 to 1.21) | 0.444 |
|  | 4 (least deprived, ref) | 1.00 | - | 1.00 | - |
| Occupation | Self-employed | 1.73 (1.45 to 2.06) | <0.001 | 1.86 (1.56 to 2.21) | <0.001 |
|  | Retired | 0.63 (0.55 to 0.72) | 0.031 | 0.85 (0.72 to 0.99) | 0.033 |
|  | Furloughed | 2.18 (1.60 to 2.97) | <0.001 | 2.18 (1.60 to 2.97) | <0.001 |
|  | Unemployed | 7.76 (5.50 to 11.0) | <0.001 | 7.76 (5.50 to 11.0) | <0.001 |
|  | Student | 1.15 (0.81 to 1.64) | 0.558 | 1.11 (0.78 to 1.59) | 0.549 |
|  | Other/never employed/sick | 2.08 (1.59 to 2.62) | <0.001 | 2.10 (1.63 to 2.69) | <0.001 |
|  | Employed (ref) | 1.00 | - | 1.00 | - |
| Housing | Mortgage | 1.66 (1.45 to 1.89) | <0.001 | 1.53 (1.34 to 1.74) | <0.001 |
|  | Private Renting | 4.55 (3.75 to 5.53) | <0.001 | 4.35 (3.58 to 5.28) | <0.001 |
|  | Renting Council | 11.6 (8.81 to 15.30) | <0.001 | 11.5 (8.72 to 15.10) | <0.001 |
|  | Other | 2.94 (2.28 to 3.79) | <0.001 | 2.77 (2.15 to 3.57) | <0.001 |
|  | Owns home (ref) | 1.00 | - | 1.00 | - |
| Self-reported general health | Poor | 5.32 (3.94 to 7.18) | <0.001 | 5.37 (3.98 to 7.26) | <0.001 |
|  | Fair | 3.41 (2.84 to 4.09) | <0.001 | 3.46 (2.88 to 4.14) | <0.001 |
|  | Good | 1.98 (1.72 to 2.29) | <0.001 | 2.01 (1.74 to 2.33) | <0.001 |
|  | Very good | 1.21 (1.06 to 1.39) | 0.003 | 1.23 (1.07 to 1.40) | 0.003 |
|  | Excellent (ref) | 1.00 | - | 1.00 | - |

To explore these findings further, we investigated whether associations between incident COVID-19 and income insufficiency were stronger for the subset of participants who either reported ‘long COVID’ or who were hospitalised for COVID-19 treatment. Results are shown in Table 3: point estimates for adjusted ORs were higher for those who reported ‘long COVID’ or hospitalisation than for those who did not, both in the short term (P values for trend 0.002 for both ‘long COVID’ and hospitalisation) and in the long term (P values for trend 0.01 and 0.03 for ‘long COVID’ and hospitalisation, respectively).

**Table 3:** Impact of self-reported ‘long COVID’ and hospitalisation for COVID-19 on reporting insufficient income during follow-up.

| Variable | Response | Acute Responses |  | Long-term Responses |  |
| --- | --- | --- | --- | --- | --- |
|  |  | Adjusted OR (95% CI) | P | Adjusted OR (95% CI) | P |
| Self-report 'long COVID' | No COVID-19 (ref) | 1.00 | - | 1.00 | - |
|  | COVID-19, no 'long COVID' | 1.44 (1.15 to 1.80) | 0.003 | 1.07 (0.91 to 1.25) | 0.445 |
|  | 'Long COVID' | 1.50 (1.14 to 1.95) | 0.002 | 1.39 (1.10 to 1.77) | 0.006 |
|  | P for trend | - | 0.002 | - | 0.011 |
| Hospitalisation due to COVID-29 | No COVID-19 (ref) | 1.00 | - | 1.00 | - |
|  | COVID-19, not hospitalised | 1.37 (1.10 to 1.71) | 0.002 | 1.14 (0.97 to 1.32) | 0.077 |
|  | COVID-19, hospitalised | 1.91 (0.694 to 5.25) | 0.220 | 1.72 (0.87 to 3.40) | 0.118 |
|  | P for trend | - | 0.002 | - | 0.026 |

*2 Multivariable regression models fully adjusted for the following baseline variables: sex, age, ethnicity, country, IMD quartile, occupation, housing and self-reported general health.*

Finally, we examined whether incident COVID-19 was associated with our secondary outcome of absence from work due to sickness. Results are presented in Table 4: incident COVID-19 was associated with increased odds of reporting sickness absence from work in the long-term (aOR 5.29, 95% CI 2.76 to 10.10) but not acutely (aOR 1.34, 95% CI 0.52 to 3.49).

**Table 4:**
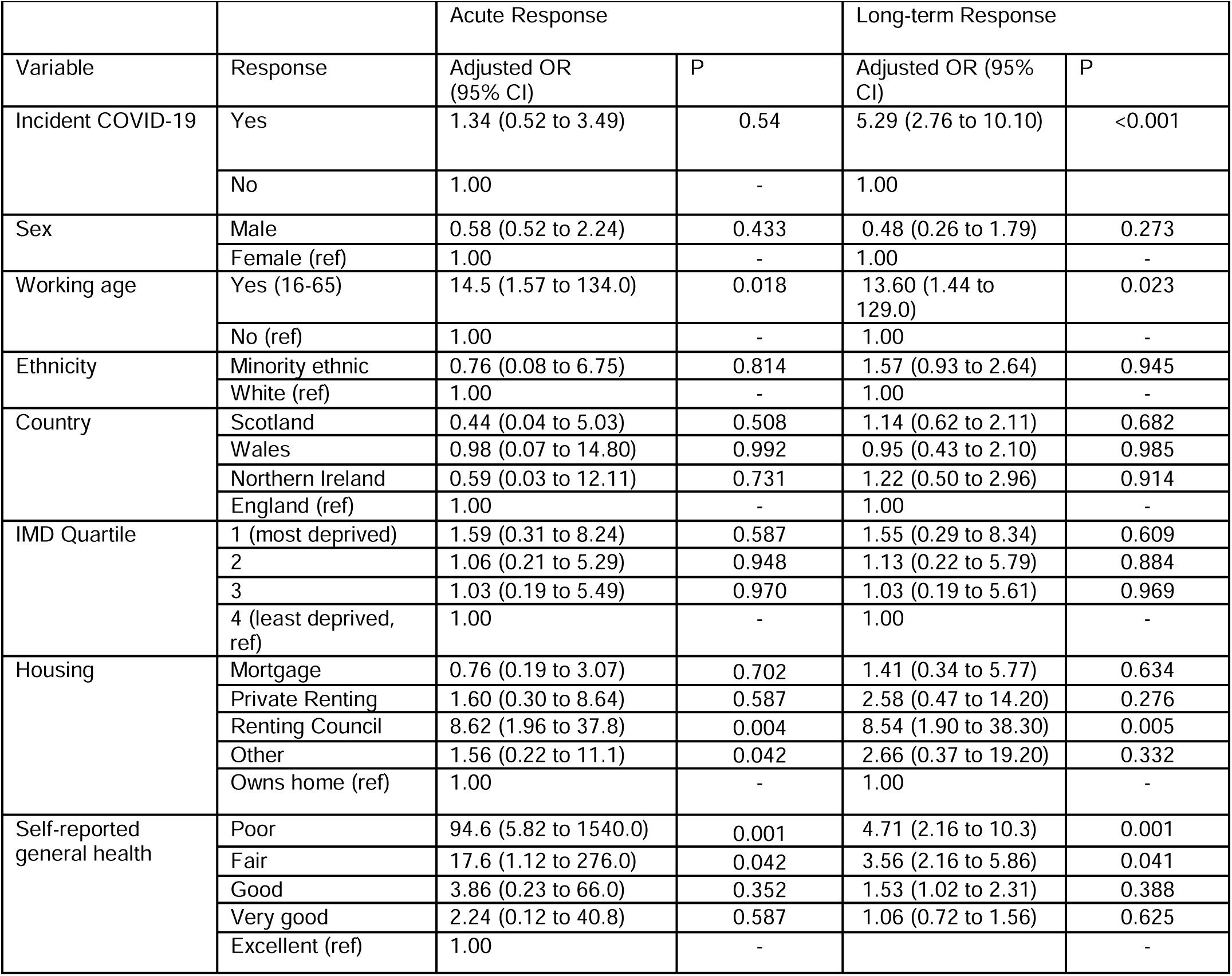
Determinants of reporting ‘not working due to sickness’ during follow-up.

| Variable | Response | Acute Response |  | Long-term Response |  |
| --- | --- | --- | --- | --- | --- |
|  |  | Adjusted OR (95% CI) | P | Adjusted OR (95% CI) | P |
| Incident COVID-19 | Yes | 1.34 (0.52 to 3.49) | 0.54 | 5.29 (2.76 to 10.10) | <0.001 |
|  | No | 1.00 | - | 1.00 |  |
| Sex | Male | 0.58 (0.52 to 2.24) | 0.433 | 0.48 (0.26 to 1.79) | 0.273 |
|  | Female (ref) | 1.00 | - | 1.00 | - |
| Working age | Yes (16-65) | 14.5 (1.57 to 134.0) | 0.018 | 13.60 (1.44 to 129.0) | 0.023 |
|  | No (ref) | 1.00 | - | 1.00 | - |
| Ethnicity | Minority ethnic | 0.76 (0.08 to 6.75) | 0.814 | 1.57 (0.93 to 2.64) | 0.945 |
|  | White (ref) | 1.00 | - | 1.00 | - |
| Country | Scotland | 0.44 (0.04 to 5.03) | 0.508 | 1.14 (0.62 to 2.11) | 0.682 |
|  | Wales | 0.98 (0.07 to 14.80) | 0.992 | 0.95 (0.43 to 2.10) | 0.985 |
|  | Northern Ireland | 0.59 (0.03 to 12.11) | 0.731 | 1.22 (0.50 to 2.96) | 0.914 |
|  | England (ref) | 1.00 | - | 1.00 | - |
| IMD Quartile | 1 (most deprived) | 1.59 (0.31 to 8.24) | 0.587 | 1.55 (0.29 to 8.34) | 0.609 |
|  | 2 | 1.06 (0.21 to 5.29) | 0.948 | 1.13 (0.22 to 5.79) | 0.884 |
|  | 3 | 1.03 (0.19 to 5.49) | 0.970 | 1.03 (0.19 to 5.61) | 0.969 |
|  | 4 (least deprived, ref) | 1.00 | - | 1.00 | - |
| Housing | Mortgage | 0.76 (0.19 to 3.07) | 0.702 | 1.41 (0.34 to 5.77) | 0.634 |
|  | Private Renting | 1.60 (0.30 to 8.64) | 0.587 | 2.58 (0.47 to 14.20) | 0.276 |
|  | Renting Council | 8.62 (1.96 to 37.8) | 0.004 | 8.54 (1.90 to 38.30) | 0.005 |
|  | Other | 1.56 (0.22 to 11.1) | 0.042 | 2.66 (0.37 to 19.20) | 0.332 |
|  | Owns home (ref) | 1.00 | - | 1.00 | - |
| Self-reported general health | Poor | 94.6 (5.82 to 1540.0) | 0.001 | 4.71 (2.16 to 10.3) | 0.001 |
|  | Fair | 17.6 (1.12 to 276.0) | 0.042 | 3.56 (2.16 to 5.86) | 0.041 |
|  | Good | 3.86 (0.23 to 66.0) | 0.352 | 1.53 (1.02 to 2.31) | 0.388 |
|  | Very good | 2.24 (0.12 to 40.8) | 0.587 | 1.06 (0.72 to 1.56) | 0.625 |
|  | Excellent (ref) | 1.00 | - |  | - |

## Discussion

To our knowledge, this study is the first to investigate the impact of COVID-19 on subsequent risk of becoming economically vulnerable. We report that incident COVID-19 was independently associated with increased risk of reporting insufficient household income, both in the short- and the long-term. Associations were stronger where COVID-19 precipitated ‘long COVID’ or hospitalisation, supporting causal interpretation. Incident COVID-19 was also associated with increased risk of reporting absence from work due to sickness in the long-term.

Our findings accord with those of studies that have investigated the impact of other infectious diseases on economic outcomes. People living with HIV have been reported to experience higher rates of severe poverty, employment loss and impaired physical and mental functioning.^13,14,15^ Similar analyses revealed a link between tuberculosis and increased poverty in both the UK and India.^16,17^ However, these studies were all cross-sectional in design, leaving uncertainty as to whether the diseases in question were a cause or consequence of the observed poverty.

Our analysis aimed to identify whether there is evidence of an association between these outcomes in a specific direction of causality, i.e. from disease to economic vulnerability. The prospective design employed in the current study was valuable to this end, as it allowed us to clearly establish the temporality of the relationship between incident COVID-19 and subsequent economic vulnerability. Demonstration of a dose-response relationship between severity of COVID-19 and the primary outcome, along with consistency of association for two different measures of economic vulnerability (inadequate income and sickness absence) both strengthen the case for causal interpretation.^18^

Taking these findings together with other research showing that socio-economic disadvantage increases the risk of developing COVID-19,^1,2,3,4^ our current study represents an important advance by indicating that the relationship between COVID-19 and socio-economic deprivation may be bi-directional. This suggests a ‘vicious cycle’ of poor health and economic vulnerability which individuals could be pushed into, or accelerated along, by COVID-19. It is notable that incident COVID-19 had a significant negative impact on self-assessed adequacy of household income both acutely and in the long-term, whereas the impact on work absence due to sickness was only evident in the long-term. One potential reason for this is that those acutely ill with COVID-19 would still self-classify as ‘employed’ but on temporary leave, whilst persistent COVID-19 symptoms might lead to a change in status to official sickness absence. This raises the possibility that COVID-19 may impact economic vulnerability through multiple mechanisms including non-employment-based mechanisms in the short term, such as increased health-related costs, as well as employment-based mechanisms in the longer term.

Our study has several strengths. Its large size afforded ample power to detect potential impacts of COVID-19 on our primary and secondary outcomes, while its population-based prospective design maximises generalisability of our findings while allowing us to characterise temporal relationships between exposures and outcomes. Detailed characterisation of participants allowed us to adjust for multiple potential confounding factors in multivariable analyses, and to explore two different indicators of economic vulnerability.

This work also has limitations. First, the variables of interest are all self-reported, including both SARS-CoV-2 test results and indicators of economic vulnerability. Participants were unaware of the hypotheses tested in this work, however, reducing potential for reporter bias to operate. Second, the study population was not perfectly representative of the adult UK population as a whole: males, younger people, people of minority ethnic origin and those with lower educational attainment were all under-represented. Further, internet access was a prerequisite to take part, which could limit generalisability of results particularly amongst the most economically deprived. While this may have limited our power to detect associations within sub-groups, we highlight that representativeness is not necessarily a barrier to identification of causal associations in observational epidemiology.^19^ Third, as with any observational study, residual or unmeasured confounding cannot be ruled out as an explanation for the associations we observe. Finally, we handled missing data under the assumption that survey data were missing at random. It is possible that data were more likely to be missing if someone had COVID-19 or became economically vulnerable. In the most extreme case, fatal or very severe COVID-19 would prevent questionnaire completion; alternatively, someone may have become ill or lost their job then no longer have the cognitive or physical capacity to complete the questionnaires. Conversely, it is possible that SARS-CoV-2 test positivity may have increased the likelihood of participants completing their monthly follow-up questionnaires.

Our findings highlight the need for further research in three areas. First, analogous studies should be done in other populations to determine whether our findings can be replicated; ideally such studies should capture details of longitudinal earnings to introduce greater objectivity and quantification of impacts while reducing reporting bias. Second, further work is needed to understand the specific mechanisms by which COVID-19 may lead to economic vulnerability, investigating the relative importance of factors including lost employment, ‘long COVID’ symptoms and stigmatisation. Third, our findings suggest the need for further work to explore bi-directional relationships between illness and deprivation more generally.

In conclusion, we report independent associations between incident COVID-19 and subsequent development of economic vulnerability, exposing a previously hidden human cost of the pandemic. Our findings have potentially significant policy implications, given the economic imperative to plan COVID-19-related spending in the most efficient way possible. While a ‘vicious cycle’ of sickness and poverty presents a major threat to wellbeing, its recognition could also offer an opportunity for effective, early-stage circuit-breaker interventions with potential to avert greater costs in the future.

## Supporting information

STROBE checklist

## Data Availability

De-identified participant data will be made available upon reasonable request to Prof Martineau, subject to terms of Research Ethics Committee approval and Sponsor requirements.

