## Supplementary files for "Acute and long-term impacts of COVID-19 on economic vulnerability: a population-based longitudinal study (COVIDENCE UK)"

**Supplementary Material**

**Table S1: Baseline questionnaire**

| **Sociodemographic** | |
| --- | --- |
| Date of birth (DD/MM/YYYY) |  |
| Date of questionnaire (DD/MM/YYYY) |  |
| Post code |  |
| Please state your **assigned sex at birth.** | -Male  -Female |
| What is your ethnic origin? | - White   - English / Welsh / Scottish / Northern Irish / British - Irish - Gypsy or Irish Traveller - Any other white background   - Mixed / Multiple ethnic groups   - White and Black Caribbean - White and Black African - White and Asian - Any other Mixed / Multiple ethnic backgrounds   - Asian / Asian British   - Indian - Pakistani - Bangladeshi - Chinese - Any other Asian background   - Black / African / Caribbean / Black British   - African - Caribbean - Any other Black / African / Caribbean background   - Arab  - Other Ethnic Group |
| In the last month, was your household income sufficient to cover the basic needs of your household, such as food and heating? | - Yes  - Mostly  - Sometimes  - No |
| Please select the box that best describes your current housing situation: | - I own my home outright  - I own my home and I am paying a mortgage  - I am renting privately  - I am renting from the council/housing association  - I am staying with friends or family  - I am homeless or living in temporary accommodation  - Other |
| Do you currently claim Universal Credit? | - Yes, I have applied to receive Universal Credit but have **not yet** received any payments  - Yes, I have claimed Universal Credit and received **one or more** payments  - No |
| Which of the following best describes your current occupational status? | - Employed  - Self-employed  - Retired  - Furloughed  - Unemployed  - Student  - Never employed  - Not working due to sickness/disability or illness  - Other |
| Over the last 12 months, would you say that on the whole, your health has been: | - Excellent  - Very good  - Good  - Fair  - Poor |
| Since February 1st 2020, have you had a nose/throat swab to test for COVID-19? | - Yes  - No |
| On what date did you have this nose/throat swab?  If you are not sure of the exact date, enter the approximate date (DD/MM/YYYY).  *e.g. 25/04/2020* |  |
| What was the result? | - Positive  - Negative  - Not known |

**Table S2: Monthly follow-up questionnaire**

| **Questions asked at every monthly follow-up** |  |
| --- | --- |
| Since you last checked in with us, have you had a nose or throat swab for COVID-19 or any other respiratory virus, or has a result from a previous swab test become newly available?(This question is about tests to detect the virus itself: they are usually done in somebody who has symptoms, but screening of asymptomatic people can also be done. It’s usually a nose/throat swab, but saliva tests are also becoming available) | - Yes  - No |
| On what date did you have this nose / throat swab? If you are not sure of the exact date, enter the approximate date (DD/MM/YYYY). |  |
| What was the result? Click as many as apply. | - Positive for COVID-19 (SARS-CoV-2 coronavirus)  - Positive for influenza virus  - Positive for another respiratory virus  - Negative for all/any viruses tested  - Not Known |
| Did you go to hospital for treatment of these symptoms? | - Yes, and I was admitted to hospital (i.e. I spent one or more nights as a hospital in-patient)  - Yes, I attended a hospital accident and emergency department but I was not admitted to hospital (i.e. I went home without spending one or more nights as a hospital in-patient)  - No, I didn’t go to hospital for treatment of these symptoms |
| What did the hospital doctors diagnose? | - Suspected or proven COVID-19  - Something else |
| Would YOU say that you currently have 'long COVID', i.e. ongoing symptoms more than four weeks after the onset of proven or suspected COVID-19? | - Yes  - No  - Don’t know/not sure |
| **Since you last checked in with us,** has your household income been sufficient to cover the basic needs of your household, such as food and heating? | - Yes  - Mostly  - Sometimes  - No |
| Has your employment status changed since you last checked in with us? | - Yes  - No |
| Which of the following best describes your current occupational status? | - Employed  - Self-employed  - Retired  - Furloughed  - Unemployed  - Student  - Never employed  - Not working due to sickness/disability or illness  - Other |

**Table 3: Summary of participant response numbers for monthly-varying independent and dependent variables**

|  | Reponses | Number Participants (%) |
| --- | --- | --- |
| Dependent Variables | | |
| Income sufficient | No | 7310 (43.3%) |
|  | Yes | 9600 (64.7%) |
| Not working due to sickness | Yes | 398 (2.4%) |
|  | No | 16,512 (97.6%) |
| Independent Variables | | |
| Incident COVID-19 | Yes | 1120 (6.6%) |
|  | No | 15,790 (93.4%) |
| Self-reported ‘long COVID’ | ‘Long COVID’ | 308 (1.8%) |
|  | COVID-19, never ‘long COVID’ | 812 (4.8%) |
|  | No COVID-19 | 15,790 (93.4%) |
| Hospitalisation due to COVID-19 | COVID-19, hospitalised | 39 (0.2%) |
|  | COVID-19, never hospitalised | 1089 (6.5%) |
|  | No COVID-19 (ref) | 15,782 (93.3%) |

1Participant numbers are reported here based on whether they EVER report the answer of interest during follow-up. For income sufficiency, they are coded as ‘no’ if they answer ‘no’, ‘sometimes’ or ‘mostly’ at least once during follow-up. For sickness absence, incident COVID-19, ‘long COVID’, and hospitalisation, they are coded as ‘yes’ if they ‘yes’ for the relevant variable at least once during follow-up.

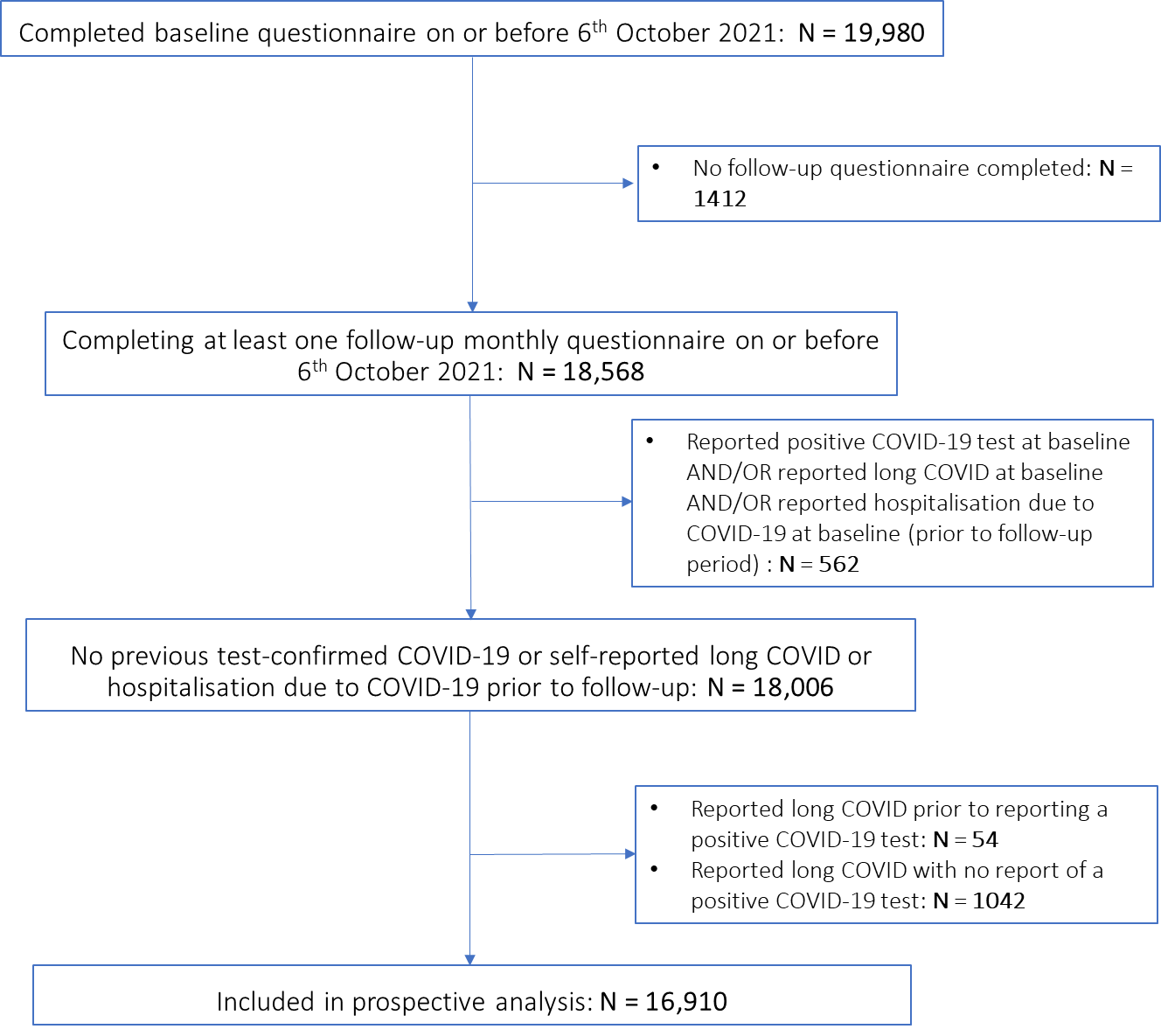
**Figure S1: Participant flow diagram**
